# Whole-exome Sequencing Study of Hypospadias

**DOI:** 10.1101/2022.01.19.22269564

**Authors:** Zhongzhong Chen, Yunping Lei, Richard H. Finnell, Zhixi Su, Yaping Wang, Hua Xie, Fang Chen

## Abstract

While hypospadias is one of the most common male congenital disorders, the missing heritability contributed by rare variants with larger effects is poorly understood. To systematically explore the variant patterns in the developing of hypospadias, we performed whole exome sequencing (WES) in 191 severe hypospadias cohort and three trios. Subsequent RNA sequencing of 12 severe hypospadiac foreskins and 6 non-hypospadiac foreskins were conducted. Among previous reported hypospadias risk associated genes, we found that *NR5A1, SRD5A2* and *AR* genes are mutational hotspots in the etiology of severe hypospadias. Additionally, rare damaging variants in novel identified outer dynein arm heavy chain (*ODNAH*) genes (*DNAH5, DNAH8, DNAH9, DNAH11*, and *DNAH17*) (*p* = 4.8×10^−17^) were significantly enriched in 191 sporadic severe hypospadias compared with 208 controls. The following transcriptomic analysis further demonstrated that the mutations in the *DNAH8* and *DNAH17* genes might affect the network regulation of testosterone (T)-dihydrotestosterone-androgen receptor (T-DHT-AR) signaling. We also identified a novel rare damaging variant of *DNAH8* in a severe hypospadias case which was transmitted from the mother. Overall, a panel of *ODNAH* genes with rare damaging variants were identified in 22.5% of severe hypospadias patients. This study provides unequivocal evidence for association of *ODNAH* genes and hypospadias. This knowledge may guide the genetic counseling for hypospadias.

**One Sentence Summary:** Rare damaging variants in outer DNAH genes were identified in 22.5% of severe hypospadias patients, which may guide the genetic counseling of families facing familial hypospadias.

## INTRODUCTION

Hypospadias is one of the most common congenital disorders in the male reproductive system. It occurs in approximately 2 per 1000 births, and its prevalence is increasing during 1980–2010^1^. Hypospadias is considered to be a complex congenital disorder caused by multiple genetic and environmental interacting factors^2^, with heritability ranged from 57% to 77%^3,4^. Paternal subfertility, 24% of which may lead to hypospadiac cases in a blind case-control study^5^, is one of the multiple risk factors responsible for the development of hypospadias. However, the genetic etiology of hypospadias is still unclear.

The penile development is controlled by AR in the prenatal period, and mediated by the balance between androgen receptor (AR) and estrogen receptor α (ESR1) levels in the neonatal period^6^. Recently, Chen et al.^7^ demonstrated that AR and ESR1 signaling play an important role in the development of mild and severe hypospadias. There are ∼30 genes in multiple pathways which are associated with the etiology of hypospadias, including: WNT signaling pathway, BMP signaling pathway, FGF signaling pathway, HH (Hedgehog) signaling pathway, Homeobox genes and others^8^. Interestingly, 79% (22/28) of previous reported hypospadias risk associated genes directly or indirectly interacted with AR and ESR1^9^. Risk associated polymorphisms with hypospadias were found in *AR, ESR1, ESR2, DGKK, SRD5A2, CYP1A1, SD17B3, FGF8, FGFR2, HMID1, ATF3, MAMLD1, GSTM1* and *GSTT1*^10^. Additionally, two large genome-wide association studies (GWAS) cohorts for hypospadias have found ∼24 susceptibility loci^11,12^. Subsequently, Kojima et al.^13^ identified SNPs in *HAAO* and *IRX6* genes were associated with hypospadias in Japanese population. And Chen et al. identified rs11170516, within the *SP1*/*SP7*, was associated with moderate-severe hypospadias^9^. However, the GWASs only explained about 9.4% of the genetic variance^11,12^, and the majority of genetic contribution of hypospadias remains unknown.

In human hypospadias, the majority of genetic studies have either been variant analysis in GWAS studies or key genes in large affected hypospadias cohorts. Recently, Kalfa team attempted to address the “missing heritability” in hypospadias using targeted Next-generation sequencing (NGS) panel including 336 genes^14^. While they are valuable, these variants still give rise to “missing heritability” and do not prove pathogenicity^15^. Previous investigations demonstrated that missense and loss-of-function (LoF) variants are more likely than other variant types to adversely affect protein function; therefore, they are considered more likely to be causative than other types of genetic variants^16^. Rare damaging variants (LoF and deleterious missense variants), which dramatically alter protein sequences, may explain the “missing heritability” missed by GWAS analysis, and were used in identifying causal genes in birth defect diseases, such as neural tube defects^17,18^ and congenital heart disease^19^. These studies suggest that the rare damaging variants may contribute to the etiology of hypospadias in humans; however, there is a paucity of such data on human hypospadias. A more extensive genetic panel as well as statistical evidence are required to explore the genetic etiology of hypospadias^15^.

Unlike most previous hypospadias studies that focused primarily on common variant associations^11,12^ or candidate genes^14^, we performed whole exome sequencing (WES) and RNA sequencing in a large hypospadias cohort. We identified a significant enrichment of damaging variants in outer dynein arm heavy chain (*ODNAH*) genes (*ODNAH5, ODNAH8, ODNAH9, ODNAH11, ODNAH17*) in severe hypospadias cases. This study complements the known common hypospadias risk-associated variants and indicates the role of rare damaging variants in hypospadias development.

## METHODS

### Ethical compliance

The study was conducted with approval of the Ethics Committee of the Shanghai Children’s Hospital in China (approve #: 2020R018-E01). Patients with hypospadias were diagnosed by the Department of Urology. Patients with congenital penile curvature but without hypospadias were excluded from this study. Each patient was informed of the purpose of this study, and written consent forms were obtained from all participants or their parent/legal guardian. Ethical approval was obtained from the Shanghai Children’s Hospital in China.

### Human Subjects

We enrolled 194 patients (mean age 2.04 ± 0.09 years) undergoing repair of severe hypospadias from 2011 to 2019 at Shanghai Children’s Hospital, Shanghai Jiao Tong University. We performed whole exome sequencing (WES) on 200 samples (including three trios) and RNA sequencing on 18 samples.

For RNA sequencing analysis, 18 prepuces of children who underwent consecutive circumcision either because of phimosis (controls; n = 6; mean age 2.84 ± 0.22 years) or because of hypospadias repair (*AR* mutated severe hypospadias, n = 4; *DNAH8* mutated severe hypospadias, n = 4; *DNAH17* mutated severe hypospadias, n = 4), were included in this study.

### Rare variants and functional annotation

In this study, we initially systematically investigated the mutation patterns of rare variants in a Chinese population composed of 191 hypospadias and 208 Chinese Han population (CHS + CHB) from 1KGP (1000 Genomes Project). We performed the WES analysis and functional annotation using the method of Chen^18^. As described in this method, coding variants were further classified as LoF (loss of function, including splice acceptor/donor, stop gained/lost, initiator codon and frameshift indels), missense and others. The missense variants, that were predicted to be deleterious by Sorting Intolerant From Tolerant (SIFT)^20^ and damaging by Polymorphism Phenotyping version 2 (PolyPhen-2)^21^, were annotated as deleterious missense variants (D-mis). Rare variants with minor allele frequency (MAF) > 1% in hypospadias or controls cohort were excluded. Rare damaging variants (LoF and D-mis) were further selected that had a MAF < 0.1% in 1KGP and ExAC databases (http://exac.broadinstitute.org). The rare damaging variants in 30 previous reported hypospadias risk associated genes^8,9^, including *SHH, GLI1, GLI2, GLI3, FGF8, FGF10, FGFR2, BMP7, WT1, DGKK, HSD3B1, HSD17B3, SRD5A2, AR, ESR1, ESR2, BMP4, HOXA4, HOXB6, MAP3K1, CHD7, NR5A1, MAMLD1, HSD3B2, CYP11A1, AKR1C3, ATF3, BNC2, SP1* and *SP7*, were identified to demonstrate their genetic contribution to hypospadias. Twenty-seven variants in *DNAH8, DNAH9* and *DNAH17*, identified by WES and predicted likely to be damaging, were selected to be further validated by Sanger sequencing. *DNAH* gene family (accession TF316836) members, including *DNAH1, DNAH2, DNAH3, DNAH5, DNAH6, DNAH7, DNAH8, DNAH9, DNAH10, DNAH11, DNAH12, DNAH14* and *DNAH17*, were identified using the curated phylogenetic database of EMBL-EBI (http://www.treefam.org/family/TF316836).

### RNA sequencing and differential gene expression analysis

The RNA was extracted from human tissues with TRIzol reagent according to the manufacturer’s instructions (Life Technologies; CA, USA). RNA samples that pass quality parameters were then used to develop RNA libraries using TruSeq RNA Library Prep Kit v2 (Illumina, San Diego, CA, USA) following the standard manufacturer’s protocol. The libraries were sequenced on Illumina NovaSeq 6000 platform with 150 bp paired-end sequencing.

RNA-Seq fastq files were generated from the Illumina NovaSeq 6000 sequencer. Skewer software^22^ was then used to dynamically remove the 3’ ends, linker sequences, and low mass fragments of raw reads. Quality assessment was carried out by FastQC tool (www.bioinformatics.babraham.ac.uk/projects/fastqc/). Trimmed clean reads were then mapped to the human reference genome (GRCh38) using STAR^23^. Then, the transcriptome was assembled via StringTie software^24^ based on the Ensembl database^25^ annotation. The FPKM (Fragments Per Kilobase of transcript per Million mapped reads) method was applied to assess differentially expressed genes. All statistical tests were conducted using two-tailed Student’s t tests with *p* < 0.05 as the significance level. Function heatmap.2 was conducted for the graphical display of the dendrogram^26^. GeneSense^27^ and STRING (https://www.string-db.org) were used to identify protein-protein interactions (PPIs) encoded by hypospadias risk associated genes in human. All statistical analyses were performed by R software (http://www.R-project.org).

RNA Expression data of 27 different tissues from 95 individuals, which is part of the Human Protein Atlas (www.proteinatlas.org) (BioProject: PRJEB4337)^28^, was taken from NCBI database.

## RESULTS

### Genetic spectrum of rare damaging variants in known hypospadias risk associated genes

We have sequenced the whole exome genomes (WES) of 191 severe hypospadias individuals using Illumina technology to a mean depth of 142× and achieve a 99.9% breadth of coverage of target nucleotides (Figure 1). On average, each hypospadias participant contains ∼123 rare damaging variants (Figure 1), which is close to the number of the protein-truncating variants in the UK Biobank array^29^.

**Figure 1.**
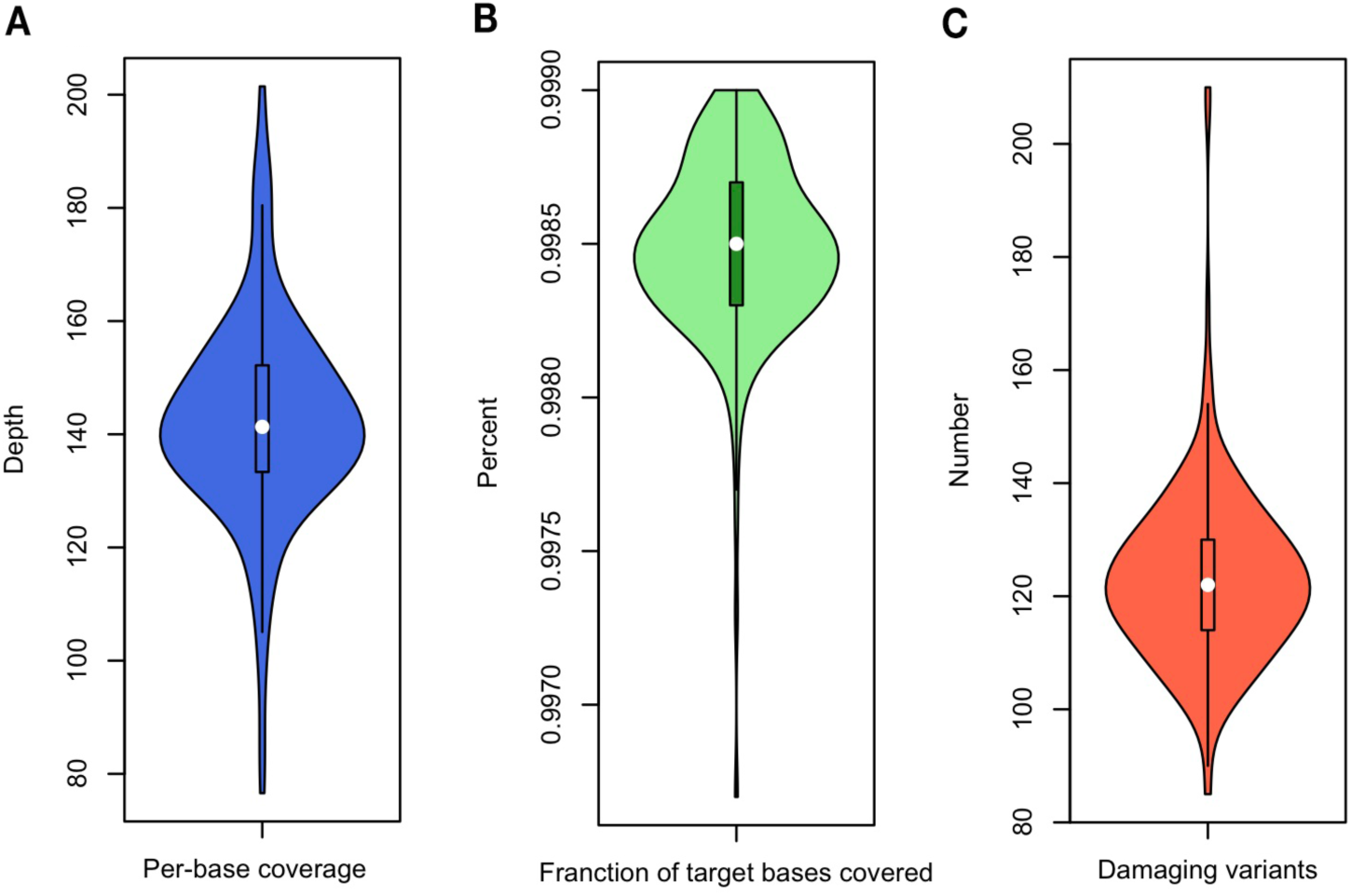
Distribution of per-base depth of coverage, fraction of target bases covered, and the number of individual’s rare damaging variants.

In this study, we investigated the rare damaging variants of 30 previous reported hypospadias risk associated genes from 191 severe hypospadias cases, which identified 27.2% (52/191) of hypospadias patients carried rare damaging variants in 21 genes, including *NR5A1, SRD5A2, AR, GLI2, HSD3B2, GLI3, AKR1C3, ESR2, FGFR2, HSD3B1, MAP3K1, BMP4, BMP7, CHD7, DGKK, ESR1, FGF8, HOXA4, SHH, SP7, WT1* (Figure 2). Among all detected 21 genes, 11 genes (*NR5A1, SRD5A2, AR, GLI2, HSD3B2, GLI3, AKR1C3, ESR2, FGFR2, HSD3B1, MAP3K1*) with rare damaging variants were observed in more than one cases. The top three genes with the highest percentage of mutations in our human severe hypospadias cohort are *NR5A1, SRD5A2* and *AR*. This included 5.2% cases with 5.2% (10/191) *NR5A1*, 5.2% (10/191) cases with *SRD5A2* and 4.2% (8/191) cases with *AR*. Therefore, rare damaging mutations of *NR5A1, SRD5A2* and *AR* genes occurs in approximately 14% of all severe hypospadias cohort. We also identified that 2.6%, 2.1% and 1.6% cases had rare damaging variants in *GLI2, HSD3B2, GLI3*, respectively. This suggested that rare damaging mutations in *AR, NR5A1* and *SRD5A2* genes function as mutational hotspots in the etiology of severe hypospadias.

**Figure 2.**
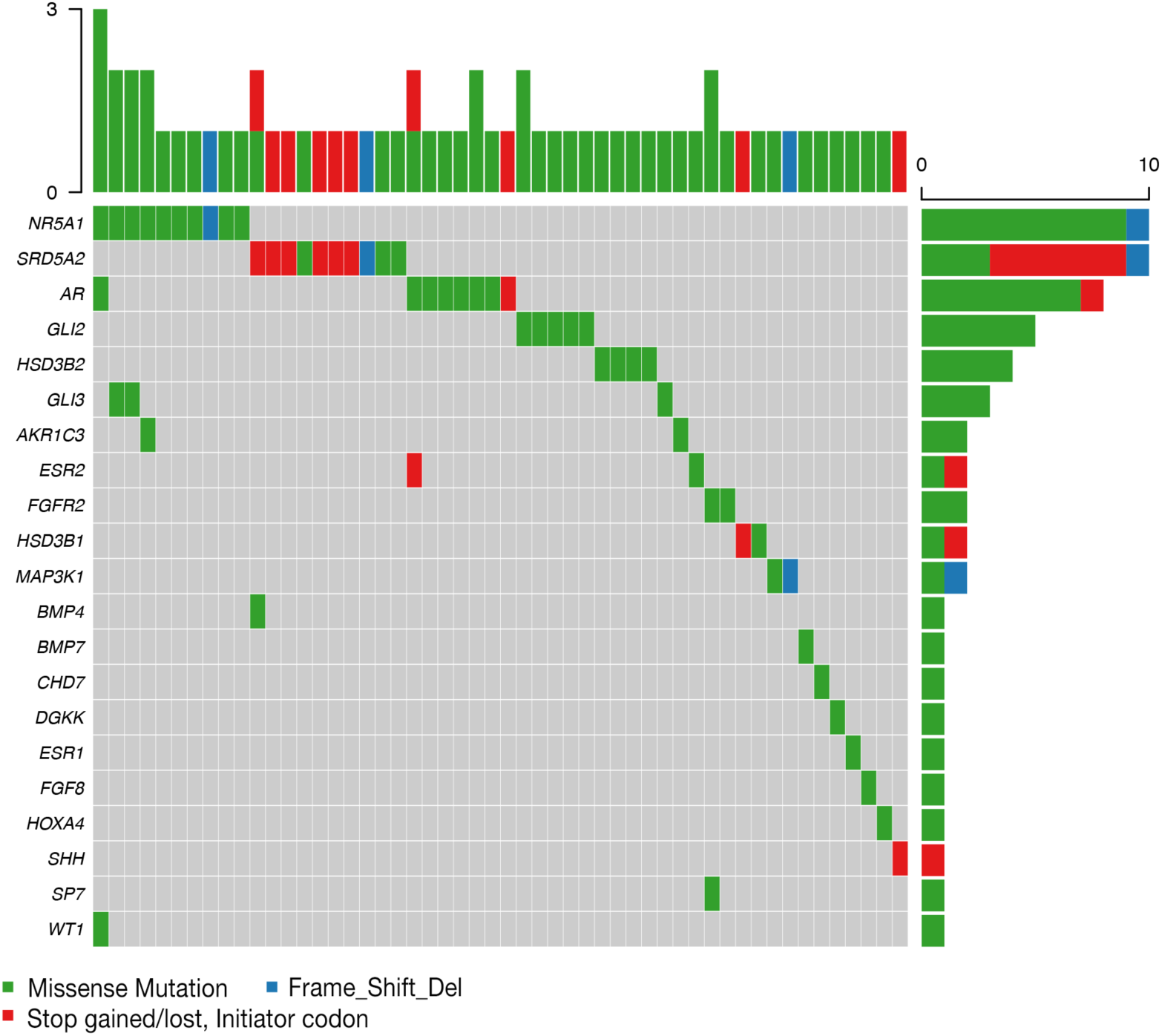
Damaging variants of 27.2% (52/191) severe hypospadias with known risk associated genes.

### Burden of genes with rare damaging variants in *ODNAH* genes

We next carried out genetic burden tests to investigate the role of rare damaging variants in the development of urethra across the genome for association with the risk of severe hypospadias. A significant increase of rare damaging variants was first identified in outer dynein arm heavy chain (*ODNAH*) genes, including *DNAH5, DNAH8, DNAH9, DNAH11* and *DNAH17*. A significant excess of rare damaging variants was identified in *ODNAH* genes in severe hypospadias (*p* = 4.8 × 10^−17^) (Table 1). *DNAH5, DNAH9* and *DNAH11*, considered motile cilia-specific expressed genes^30^, enriched more rare damaging variants than *DNAH8* and *DNAH17* genes (Table 1). Apart from *ODNAH* genes, genetic burden in inner dynein arm heavy chain (*IDNAH*) genes from *DNAH* family (TF316836) was further evaluated (Table 2). *DNAH1* and *DNAH2* were significant enrichment of rare damaging variants (*p* < 0.01), whereas no significant excess of rare damaging variants was identified in *DNAH3, DNAH6, DNAH7, DNAH10* and *DNAH12*. Compared with the rare damaging variants from controls, rare damaging variants in *ODNAH* genes were significant enriched in severe hypospadias cohort. These data demonstrate rare damaging variants of the *ODNAH* genes, identified in ∼22.5% of (43/191) sporadic hypospadias cases, are expected to contribute the genetic etiology of hypospadias.

**Table 1.** Burden analysis of rare damaging variants in axonemal outer dynein arm heavy chain (*ODNAH*) genes by severe hypospadias vs. control comparison.

| Genes | Number of rare damaging variants |  | OR | *P*<sup>a</sup> |
| --- | --- | --- | --- | --- |
|  | Cases | Controls |  |  |
| Total | 51 | 0 | NA | 4.8×10<sup>-17</sup> |
| *DNAH5* | 10 | 0 | NA | 6.3×10<sup>-4</sup> |
| *DNAH8* | 7 | 0 | NA | 5.8×10<sup>-3</sup> |
| *DNAH9* | 13 | 0 | NA | 6.9×10<sup>-5</sup> |
| *DNAH11* | 13 | 0 | NA | 6.9×10<sup>-5</sup> |
| *DNAH17* | 8 | 0 | NA | 2.8×10<sup>-3</sup> |

<sup>a</sup>*P*-value based on binomial test.

**Table 2.** Burden analysis of rare damaging variants in axonemal inner dynein arm heavy chain (*IDNAH*) genes by severe hypospadias vs. control comparison.

| Genes | Number of rare damaging variants |  | OR | *P*<sup>a</sup> |
| --- | --- | --- | --- | --- |
|  | Cases | Controls |  |  |
| *DNAH1* | 11 | 0 | NA | 3.0×10<sup>-4</sup> |
| *DNAH2* | 11 | 1 | 11.98 | 2.4×10<sup>-3</sup> |
| *DNAH3* | 23 | 15 | 1.48 | 0.14 |
| *DNAH6* | 8 | 7 | 1.24 | 0.80 |
| *DNAH7* | 19 | 8 | 2.59 | 0.02 |
| *DNAH10* | 10 | 2 | 5.45 | 0.02 |
| *DNAH12* | 6 | 7 | 0.91 | 1 |
| *DNAH14* | 13 | 9 | 1.77 | 0.27 |

<sup>a</sup>*P*-value based on binomial test.

### Dysregulated expression and dynamic network in *AR* and *ODNAH* mutated hypospadias

Rare damaging variants are expected to have stronger effects that disrupt transcription and lead to expression of abnormal protein that often cause loss of protein function or gain of function effects^31^. To explore how an understanding of *AR* and *ODNAH* genes accounts for severe hypospadias caused by rare damaging variants, 12 patients (four *AR* mutated cases, four *DNAH8* mutated cases and four *DNAH7* mutated cases) out of 191 patients and six controls were analyzed using RNA-seq. Unsupervised hierarchical cluster analysis revealed the 15 out of 30 previously reported hypospadias risk associated genes that differ more between hypospadias and controls (Figure 3A). We subsequently conducted the network propagation analysis to measure the biological regulatory influenced by rare damaging variants in *AR, DNAH8* and *DNAH17*. In *AR* mutated patients, SHH genes (*GLI1* and *GLI2*), estrogen-regulating gene *ATF3*, gonad development related gene *CHD7*, and *DNAH17* were significantly expressed compared with controls (*p* < 0.05) (Figure 3B). We investigated *AR* gene and hypospadias risk associated genes by protein-protein interactions (PPIs) analysis, indicating that rare damaging variants in *AR* may directly alter the expression *GLI1, GLI2, ATF3* and *CHD7* and indirectly affect the expression of *DNAH17* (Figure 3B). Compared to controls, *AR, DNAH17, ATF3, CHD7* and homeobox genes (*HOXA4* and *HOXB6*) were significantly expressed in patients with *DNAH8* mutations (*p* < 0.05) (Figure 3C). The PPI network connects to DNAH17 revealed that the expression of *AR, DNAH17, ATF3* and *CHD7* might be directly affected by *DNAH8*, whereas *HOXA4* and *HOXB6* might be indirectly impacted by *DNAH8* (Figure 3C). In *DNAH17* mutated patients, *AR, SRD5A2, GLI1, GLI2, GLI3, BMP4, ATF3, CHD7, HSD17B3, HOXA4, HOXB6, MAMLD1* and *COL6A3* were significantly expressed in severe hypospadias (*p* < 0.05) (Figure 3D). The PPI network with DNAH17 protein as the hub (Figure 7B) demonstrated that all genes might be indirectly affected by the rare damaging variants in *DNAH17* (Figure 3D). Additionally, *AR, ATF3, CHD7* and *DNAH17*, which act as the crucial nodes with the highest connectivity in these three subnetworks, might be closely associated with the development of hypospadias.

**Figure 3.**
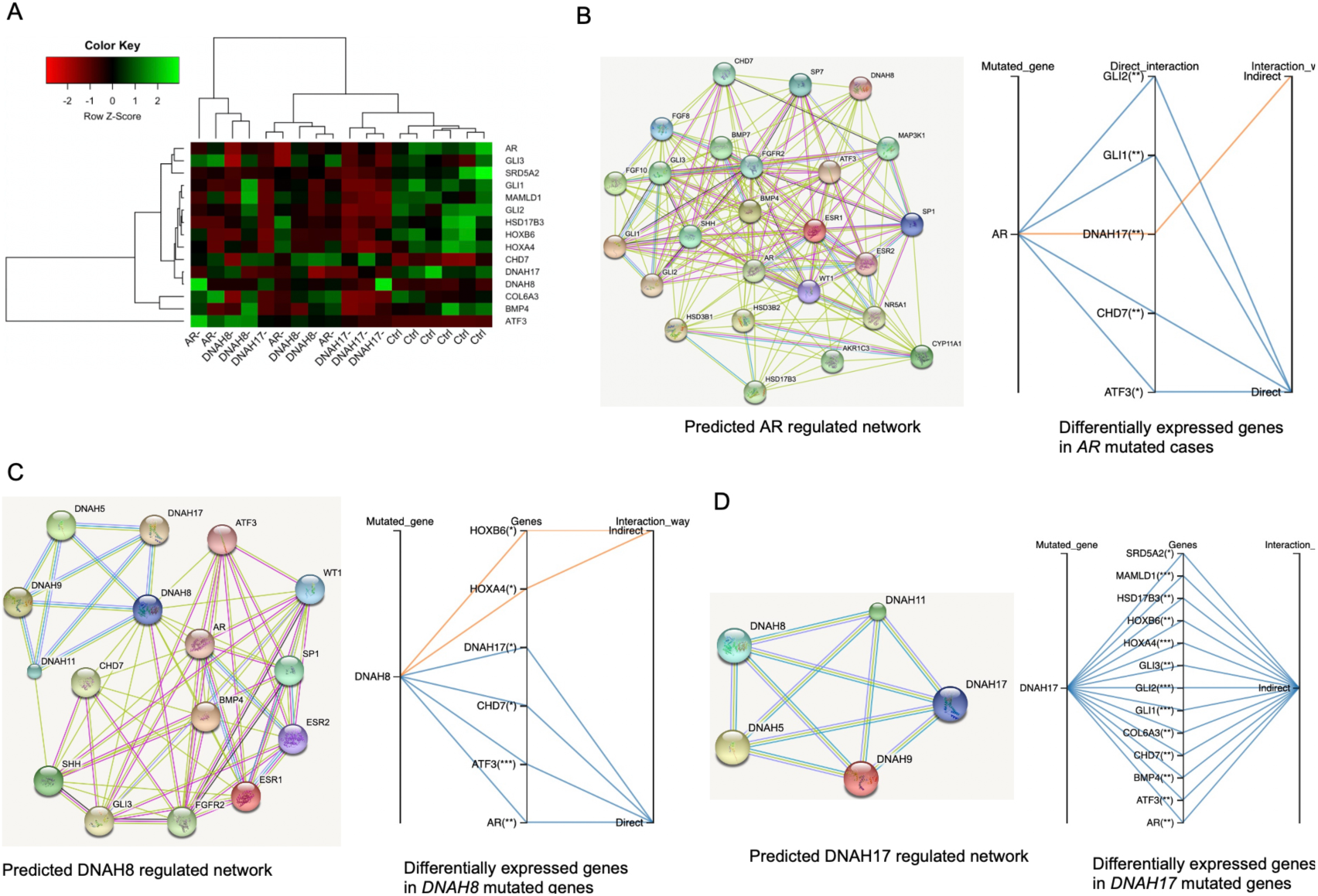
Differential mRNA expression of known hypospadias risk genes in *AR, DNAH8* and *DNAH17* mutated severe hypospadias cases compared with controls. (A) Hierarchical clustering 15 differentially expressed genes in *AR, DNAH8* and *DNAH17* mutated patients compared with controls. (B) Predicted protein-protein interactions (PPIs) network of AR, differentially expressed genes directly or indirectly interacted with AR in AR mutated cases based on PPIs. (C) PPIs network of DNAH8, differentially expressed genes directly or indirectly interacted with DNAH8 in *DNAH8* mutated cases based on PPIs. (D) PPIs network of DNAH17, differentially expressed genes directly or indirectly interacted with DNAH17 in *DNAH17* mutated cases based on PPIs.

### Potential mechanism, inheritance pattern of *ODNAH* genes in hypospadias

The different gene expression patterns demonstrated that *AR, ATF3* and *CHD7* are dysregulated in patients carried *DNAH8* and *DNAH17* mutations. Previous study indicated that *ATF3* and *CHD7* may interacted with *AR*, which is a hub gene and play a key role in the etiology of hypospadias^9^. In addition, the expression of *SRD5A2*, which is involved in the conversion of testosterone (T) to dihydrotestosterone (DHT), is significantly decreased in patients with *DNAH17* variants. Based on differential gene expression and network analysis, we proposed that *ODNAH* genes are likely to cause hypospadias by affecting the T-DHT-AR signaling, which are critical in the development of external genitalia. Furthermore, *ODNAH* genes, including *DNAH5, DNAH8, DNAH9, DNAH11* and *DNAH17* are expressed higher average mRNA levels in testis tissue compared to other 26 tissues (Figure 4A). Among these genes, *DNAH8* and *DNAH17* are exclusively highly expressed in testis tissue. *ODNAH* genes, which are specifically expressed in human testis and sex-limited, are thus expected to under differential selection. To examine the inheritance patterns that are caused by inherited or de novo mutations, we recruited three family-based trios to decipher the genetic predisposition. Of these three trios, one novel rare damaging variant, was identified in the testis-exclusive gene *DNAH8* (Figure 4B). This variant p.Pro1564Ser or p.P1564S (NM_001206927.1:c.4690C>T) in *DNAH8* gene did not exist in 1000 Genomes Project (1KGP), and very rare in in the Exome Aggregation Consortium (ExAC) database (MAF = 1.6×10^−5^). The DNAH8 1564 proline residue is located in a highly conserved region across species, and was inherited from the mother.

**Figure 4.**
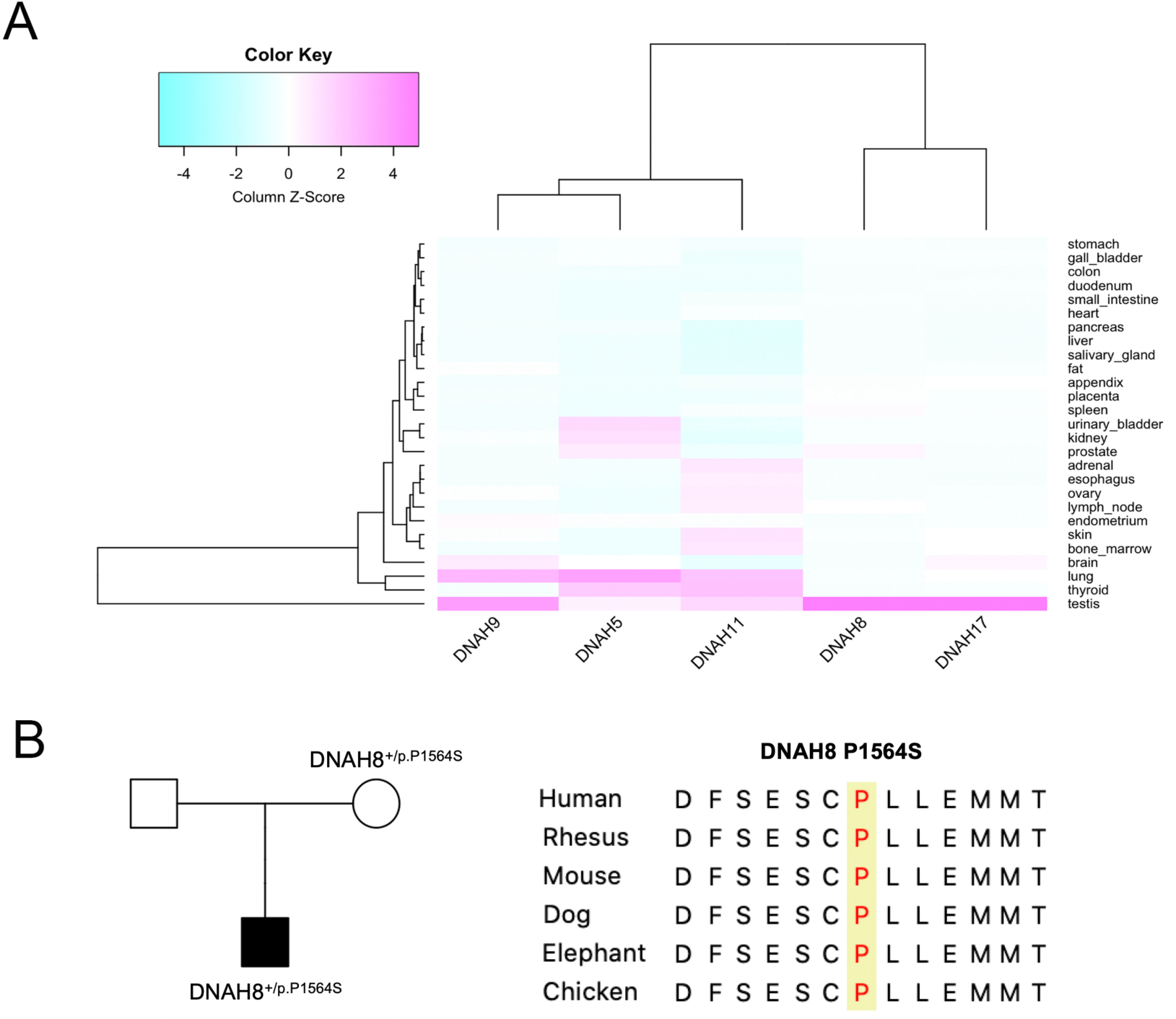
Tissue expression and pedigree analysis of *ODNAH* genes in the development of hypospadias. (A) *DNAH5, DNAH8, DNAH9, DNAH11* and *DNAH17* are highly expressed in testis compared with other tissues. (B) Pedigree of one severe hypospadias kindred with rare *DNAH8* damaging mutation is shown. The pedigree plot with shapes for male (square), female (circle). The shapes are shaded and un-shaded for hypospadias status of yes and no.

## DISCUSSION

The major heritability of hypospadias, which might be caused by subfertility, is still unknown. This is the first large-scale WES study to explore the genetic contribution of rare damaging variants and their potential regulation in a Chinese hypospadias cohort. In addition to confirming that 27.2% of severe hypospadias patients were found to carry rare damaging variants in the previously reported hypospadias risk associated genes, this study provide novel findings in several respects.

Firstly, we found that *NR5A1, SRD5A2* and *AR* genes function as mutational hotspots in the etiology of severe hypospadias. Previous studies showed that genetic causes of hypospadias accounts for about 30% of all hypospadias^32^ and 28% of severe hypospadias^33^. While the role of multiple genes and pathways are well characterized, their associations with hypospadias are not yet well understood. Given the evidence for the presence of known hypospadias associated genes^8,9^, we hypothesized that rare damaging variants may be enriched for association with risk of hypospadias. Notably, 27.2% of human severe hypospadias cohort were identified to carry rare damaging variants with reported hypospadias associated genes. Of these genes, *AR*, nuclear receptor subfamily 5 group A member 1 (*NR5A1*) and the enzyme steroid reductase 5α type 2 (*SRD5A2*) genes function as mutational hotspots in the etiology of severe hypospadias. Results showed that *AR, NR5A1* and *SRD5A2* were enriched in rare damaging variants in severe hypospadias. Among the three genes, rare damaging variants of *AR* accounted for 4.2% of severe hypospadias. The rate of this study is consistent with a previous study, which reported that mutations of *AR* accounted for 3% of hypospadias patients^34^. *NR5A1*, which encodes steroidogenic factor 1, regulates the expression of key testis genes *SOX9* and *AMH*^35^. Loss of function mutations in *NR5A1*, tend to be associated with severe hypospadias^36^. Inactivating mutations in *SRD5A2*, which converts testosterone (T) to dihydrotestosterone (DHT), can lead to a broad spectrum of masculinization defects, including hypospadias of varying phenotypes^37^. While hormone replacement therapy is an effective option for *SRD5A2* deficiency, resistance to *AR*-directed therapies are still a challenge. Taken together, mutations in *SRD5A2, AR* and *NR5A1* are crucial for understanding the genetic heterogeneity of hypospadias. Due to the possible treatment variability of this disorder, sequence analysis of *SRD5A2, AR* and *NR5A1* should be performed to better inform the diagnosis and treatment of hypospadias patients.

Another interesting discovery arising from our study is the significant enrichment of rare damaging variants of *ODNAH* genes in 22.5% severe hypospadias, indicating that *ODNAH* genes deficiency could be an important cause of hypospadias in human populations. The axoneme consists of a microtubule cytoskeleton, and is the key extracellular component of cilia and flagella in eukaryotes^38^. While it is well known that subfertility is strong correlation with hypospadias^5^, the contribution of the relevant genes is unknown. Dynein, one of the motor proteins, generate force and movement on microtubules^39^. Axonemal dyneins form the inner and outer, through ATPase activity of their heavy chains, play an important role in the beating of both cilia and sperm^40^. In this work, all five *ODNAH* genes are testis-specific genes that expressed higher average mRNA levels in testis tissue compared to other tissues (Figure 4B). The mutations in male testis-specific genes, relative to other gene groups, are more likely to morbid phenotypes, specifically in male reproduction processes^41^. Among the five *ODNAH* genes or outer dynein arms (ODAs), mutations in *DNAH17*^30,42^ and *DNAH8*^43^ have recently been reported to associated with male infertility due to asthenozoospermia, demonstrating that rare damaging variants in *ODNAH* genes tend to cause infertility. We next examined whether rare damaging variants of *IDNAH* genes or inner dynein arms (IDAs) contributed to hypospadias. Although there was no association between other *IDNAH* genes (*DNAH3, DNAH6, DNAH7, DNAH10* and *DNAH12*) and severe hypospadias, *DNAH1* and *DNAH2* showed significant enrichments of rare damaging variants (*p* < 0.01). While disruptions of *DNAH1* and *DNAH2* are associated with abnormalities of sperm flagella^44,45^, further research with larger sample sizes are warranted to delineate their genetic contributions in the development of hypospadias. In contrast to *IDNAH* genes, the significant excess of rare damaging variants in *ODNAH* genes, suggesting to induce male infertility, is therefore likely to be a major factor for hypospadiac causation.

Finally, our results demonstrated that mutations in *ODNAH* genes may be inherited from unaffected mothers and are likely to cause hypospadias by adversely affecting T-DHT-AR signaling. A previously published study has implicated AR and ESR1 signaling as having a pivotal role in the development of both mild and severe hypospadias^7^. The dysregulation of *AR* and oestrogen-responsive gene *ATF3* in *DNAH8* and *DNAH17* mutated hypospadias further provided the important support that such a link exists. Interestingly, *ODNAH* genes are highly expressed in human testes compared to other tissues (Figure 4A). The main function of the testis is to produce sperm and androgen, and T is the androgen. Mutations of *ODNAH* which specifically affect T synthesis in the testis are likely to cause hypospadias. Together with the hub role of AR in the PPI network comprising of proteins encoded by previous reported hypospadias risk associated genes^9^, we posit that *ODNAH* genes are likely to cause hypospadias by disrupting T-DHT-AR signaling. Deleterious mutations, either on viability before reproduction or on fertility, are not expected to transmitted to subsequent generations since they reduce fertility^46^. To explore the sex-limited selection, we examined three trios and identified one novel rare damaging variant of *DNAH8* transmitted from mother (Figure 4B). Consistent with the inheritance of male-infertility-causative mutations^41^, this study suggests that hypospadias-causative mutations may inherited through unaffected female, which requires further study in the future. Ongoing efforts are required to recruit more hypospadiac patients for evaluation of inheritance model.

In conclusion, this study explored the importance of rare damaging variants in hypospadias in an effort to explain the “missing heritability” identified in previously published GWAS analyses. Our studies demonstrated that identifying mutations of *NR5A1, SRD5A2* and *AR* genes are crucial for establishing an optimal treatment program for hypospadias patients. In addition, this manuscript now provides unequivocal evidence that mutations in the *ODNAH* genes (*ODNAH5, ODNAH8, ODNAH9, ODNAH11, ODNAH17*) contribute to the risk of hypospadias. This information will be informative for genetic counseling families dealing with hypospadias and male-infertility.

## Data Availability

All data produced in the present study are available upon reasonable request to the authors.

## ACKNOWLEDGMENTS

This work was supported by grants from the National Natural Science Foundation of China (81870459, 81970572).

## AUTHOR CONTRIBUTIONS

Z.C. and F.C. directed and designed the study. P.W. recruited study subjects. Z.C., X.Z. and Y.L. performed bioinformatics analysis and function annotation. Z.C., Y.L., R.F., H.X., and F.C. prepared the manuscript; all authors reviewed the manuscript.

## REFERENCES

1. Yu, X. et al. Hypospadias Prevalence and Trends in International Birth Defect Surveillance Systems, 1980-2010. Eur Urol 76, 482–490 (2019).

2. Carmichael, S.L., Shaw, G.M. & Lammer, E.J. Environmental and genetic contributors to hypospadias: a review of the epidemiologic evidence. Birth Defects Res A Clin Mol Teratol 94, 499–510 (2012).

3. Schnack, T.H. et al. Familial aggregation of hypospadias: a cohort study. Am J Epidemiol 167, 251–6 (2008).

4. Stoll, C., Alembik, Y., Roth, M.P. & Dott, B. Genetic and environmental factors in hypospadias. J Med Genet 27, 559–63 (1990).

5. Fritz, G. & Czeizel, A.E. Abnormal sperm morphology and function in the fathers of hypospadiacs. J Reprod Fertil 106, 63–6 (1996).

6. Zheng, Z., Armfield, B.A. & Cohn, M.J. Timing of androgen receptor disruption and estrogen exposure underlies a spectrum of congenital penile anomalies. Proc Natl Acad Sci U S A 112, E7194–203 (2015).

7. Chen, Z., Lin, X., Wang, Y., Xie, H. & Chen, F. Dysregulated expression of androgen metabolism genes and genetic analysis in hypospadias. Mol Genet Genomic Med 8, e1346 (2020).

8. Bouty, A., Ayers, K.L., Pask, A., Heloury, Y. & Sinclair, A.H. The Genetic and Environmental Factors Underlying Hypospadias. Sex Dev 9, 239–259 (2015).

9. Chen, Z. et al. Genome-wide association study in Chinese cohort identifies one novel hypospadias risk associated locus at 12q13.13. BMC Med Genomics 12, 196 (2019).

10. van der Zanden, L.F. et al. Aetiology of hypospadias: a systematic review of genes and environment. Hum Reprod Update 18, 260–83 (2012).

11. Geller, F. et al. Genome-wide association analyses identify variants in developmental genes associated with hypospadias. Nat Genet 46, 957–63 (2014).

12. van der Zanden, L.F. et al. Common variants in DGKK are strongly associated with risk of hypospadias. Nat Genet 43, 48–50 (2011).

13. Kojima, Y. et al. Single Nucleotide Polymorphisms of HAAO and IRX6 Genes as Risk Factors for Hypospadias. J Urol 201, 386–392 (2019).

14. Ea, V. et al. How Far Should We Explore Hypospadias? Next-generation Sequencing Applied to a Large Cohort of Hypospadiac Patients. Eur Urol (2021).

15. Chen, Z., Xie, H. & Chen, F. Re: Vuthy Ea, Anne Bergougnoux, Pascal Philibert, et al. How Far Should We Explore Hypospadias? Next-generation Sequencing Applied to a Large Cohort of Hypospadiac Patients. Eur Urol 2021;79:507–515. Eur Urol (2021).

16. Sveinbjornsson, G. et al. Weighting sequence variants based on their annotation increases power of whole-genome association studies. Nat Genet 48, 314–7 (2016).

17. Chen, Z., Kuang, L., Finnell, R.H. & Wang, H. Genetic and functional analysis of SHROOM1-4 in a Chinese neural tube defect cohort. Hum Genet 137, 195–202 (2018).

18. Chen, Z. et al. Threshold for neural tube defect risk by accumulated singleton loss-of-function variants. Cell Res 28, 1039–1041 (2018).

19. Jin, S.C. et al. Contribution of rare inherited and de novo variants in 2,871 congenital heart disease probands. Nat Genet 49, 1593–1601 (2017).

20. Kumar, P., Henikoff, S. & Ng, P.C. Predicting the effects of coding non-synonymous variants on protein function using the SIFT algorithm. Nat Protoc 4, 1073–81 (2009).

21. Adzhubei, I.A. et al. A method and server for predicting damaging missense mutations. Nat Methods 7, 248–9 (2010).

22. Jiang, H., Lei, R., Ding, S.W. & Zhu, S. Skewer: a fast and accurate adapter trimmer for next-generation sequencing paired-end reads. BMC Bioinformatics 15, 182 (2014).

23. Dobin, A. et al. STAR: ultrafast universal RNA-seq aligner. Bioinformatics 29, 15–21 (2013).

24. Pertea, M. et al. StringTie enables improved reconstruction of a transcriptome from RNA-seq reads. Nat Biotechnol 33, 290–5 (2015).

25. Cunningham, F. et al. Ensembl 2019. Nucleic Acids Res 47, D745–D751 (2019).

26. Gentleman, R.C. et al. Bioconductor: open software development for computational biology and bioinformatics. Genome Biol 5, R80 (2004).

27. Chen, Z. et al. GeneSense: a new approach for human gene annotation integrated with protein-protein interaction networks. Sci Rep 4, 4474 (2014).

28. Fagerberg, L. et al. Analysis of the human tissue-specific expression by genome-wide integration of transcriptomics and antibody-based proteomics. Mol Cell Proteomics 13, 397–406 (2014).

29. DeBoever, C. et al. Medical relevance of protein-truncating variants across 337,205 individuals in the UK Biobank study. Nat Commun 9, 1612 (2018).

30. Whitfield, M. et al. Mutations in DNAH17, Encoding a Sperm-Specific Axonemal Outer Dynein Arm Heavy Chain, Cause Isolated Male Infertility Due to Asthenozoospermia. Am J Hum Genet 105, 198–212 (2019).

31. Holbrook, J.A., Neu-Yilik, G., Hentze, M.W. & Kulozik, A.E. Nonsense-mediated decay approaches the clinic. Nat Genet 36, 801–8 (2004).

32. Sagodi, L., Kiss, A., Kiss-Toth, E. & Barkai, L. [Prevalence and possible causes of hypospadias]. Orv Hetil 155, 978–85 (2014).

33. Johnson, E.K. et al. Proximal Hypospadias-Isolated Genital Condition or Marker of More? J Urol 204, 345–352 (2020).

34. Kalfa, N. et al. Minor hypospadias: the “tip of the iceberg” of the partial androgen insensitivity syndrome. PLoS One 8, e61824 (2013).

35. Allali, S. et al. Mutation analysis of NR5A1 encoding steroidogenic factor 1 in 77 patients with 46, XY disorders of sex development (DSD) including hypospadias. PLoS One 6, e24117 (2011).

36. Kohler, B. et al. The spectrum of phenotypes associated with mutations in steroidogenic factor 1 (SF-1, NR5A1, Ad4BP) includes severe penoscrotal hypospadias in 46,XY males without adrenal insufficiency. Eur J Endocrinol 161, 237–42 (2009).

37. Chavez, B., Ramos, L., Gomez, R. & Vilchis, F. 46,XY disorder of sexual development resulting from a novel monoallelic mutation (p.Ser31Phe) in the steroid 5alpha-reductase type-2 (SRD5A2) gene. Mol Genet Genomic Med 2, 292–6 (2014).

38. Ishikawa, T. Axoneme Structure from Motile Cilia. Cold Spring Harb Perspect Biol 9(2017).

39. Roberts, A.J., Kon, T., Knight, P.J., Sutoh, K. & Burgess, S.A. Functions and mechanics of dynein motor proteins. Nat Rev Mol Cell Biol 14, 713–26 (2013).

40. King, S.M. Axonemal Dynein Arms. Cold Spring Harb Perspect Biol 8(2016).

41. Gershoni, M. & Pietrokovski, S. Reduced selection and accumulation of deleterious mutations in genes exclusively expressed in men. Nat Commun 5, 4438 (2014).

42. Zhang, B. et al. A DNAH17 missense variant causes flagella destabilization and asthenozoospermia. J Exp Med 217(2020).

43. Liu, C. et al. Bi-allelic DNAH8 Variants Lead to Multiple Morphological Abnormalities of the Sperm Flagella and Primary Male Infertility. Am J Hum Genet 107, 330–341 (2020).

44. Coutton, C. et al. Mutations in CFAP43 and CFAP44 cause male infertility and flagellum defects in Trypanosoma and human. Nat Commun 9, 686 (2018).

45. Li, Y. et al. DNAH2 is a novel candidate gene associated with multiple morphological abnormalities of the sperm flagella. Clin Genet 95, 590–600 (2019).

46. McClellan, J. & King, M.C. Genetic heterogeneity in human disease. Cell 141, 210–7 (2010).

